# Brain Dopamine Receptor System is Not Altered in Obesity: Bayesian and Frequentist Meta-Analyses

**DOI:** 10.1101/2023.06.22.23291735

**Authors:** Kyoungjune Pak, Lauri Nummenmaa

## Abstract

**Background:** Feeding induces dopamine release in the striatum, and a dysfunction of the dopaminergic reward system can lead to overeating, and obesity. Studies have reported inconsistent findings of dopamine receptor (DR) positron emission tomography (PET) scans in obesity. Here we investigated the association between DR availability and overweight/obesity using Bayesian and frequentist meta-analysis.

**Methods:** We performed a systematic search of Embase, Medline, Scopus and Web of Science for studies which compared striatal DR availability between lean subjects and overweight/obese subjects. The standardized mean difference (Hedge’s g) of DR availability was calculated after extraction of data from each study. Studies were divided into two groups according to the definition of overweight/obese subjects (BMI cutoff of 25 and 30kg/m^2^). Both Bayesian and conventional meta-analysis was done in R Statistical Software version 4.2.2 (The R Foundation for Statistical Computing).

**Results:** Nine studies were eligible for inclusion in this study. Three studies with C11-raclopride, one with C11-PNHO, two with F18-fallypride, one with I123-IBZM, one with C11-NMB and one with both C11-raclopride and C11-PNHO were included. In Bayesian meta-analysis, the standardized mean difference of DR availability between lean and overweight/obese subjects markedly overlapped with zero regardless of BMI cutoff for obesity. In frequentist meta-analysis the pooled standardized mean difference of DR availability did not show the significant difference between lean and overweight/obese subjects. There was an effect of the radiopharmaceutical on the standardized mean difference of DR availability in meta-analysis of BMI cutoff of 25 kg/m^2^.

**Conclusions:** Brain DR availability is not different between lean and overweight/obese subjects. However, the effect is dependent on the radiopharmaceutical and the degree of obesity. Further studies with multi-radiopharmaceutical in the same individuals are need to understand the association between DR and obesity.

## INTRODUCTION

Obesity has nearly tripled worldwide since 1975 and has become one of the major public health threats. Obesity is a risk factor for malignancies of the colon [1], pancreas [2], thyroid [3], liver [4], and uterus [5] as well as for cardiovascular disease and diabetes mellitus [6].

Obesity is caused by an imbalance between energy intake and expenditure over a long period of time [7]. The brain plays a critical role in controlling energy balance [7]. Feeding induces dopamine release in the striatum [8], and a dysfunction of the dopaminergic reward system can lead to overeating, and obesity [9]. However, there is no direct method to measure the synaptic dopamine concentration in the human brain. Therefore, positron emission tomography (PET) and single-photon emission computed tomography (SPECT) using dopamine receptor (DR) radiopharmaceuticals has been adopted to understand the role of dopaminergic system in obesity. A landmark study by Wang et al showed that DR availability was lower in severely obese subjects (the mean BMI of 51.2kg/m^2^) than in lean subjects [10]. After that, several studies reported inconsistent findings of DR PET scans in obesity; higher [11] or not different [12] DR availability in obese subjects compared to lean subjects. In addition, some researchers suggested non-linear relationship between DR availability and BMI [13, 14]. However, previous studies included the small sample size, the discrepancies in region-of-interest (ROI) and the variety of radiopharmaceuticals, leading to these inconsistent findings.

Therefore, it is more timely than ever to meta-analyze the previous studies of association between DR availability measured from PET or SPECT and obesity. In this study, we divided the previous publications into two groups according to the definition of overweight/obese subjects (BMI cutoff of 25 and 30kg/m^2^) and we investigated the association between DR availability and overweight/obesity with conventional as well as Bayesian meta-analysis.

## MATERIALS AND METHODS

### Data Search and Study Selection

We performed a systematic search of Embase, Medline, Scopus and Web of Science from inception to November 2022 for articles published in English using the keywords “dopamine receptor”, “obesity” and “positron emission tomography OR single-photon emission computed tomography”. All searches were limited to human studies. The inclusion criteria were neuroimaging studies which 1) compared DR availability of lean subjects and that of overweight/obese subjects and 2) measured DR availability within the striatum (whole striatum, caudate nucleus, putamen, nucleus accumbens, ventral striatum) using PET or SPECT scans. Reviews, abstracts, and editorial materials were excluded. Further, duplicate articles were excluded. If there was more than one study using the same set of patients, the study reporting information most relevant (i.e. initial patient-control differences) to the present study was included. Two authors performed the literature search and screened the articles independently, and discrepancies were resolved by consensus.

### Data Extraction and Statistical Analysis

Two reviewers independently extracted the following information from the reports: first author, year of publication, country, radiopharmaceuticals, the mean BMI of lean subjects and overweight/obese subjects and region-of-interest (ROI). First, we extracted the mean and the standard deviation of DR availability and the number of subjects in each group, directly from each study, if provided by the authors. If they were not provided by the authors, BMI and DR availability were extracted from the figures using WebPlotDigitizer version 4.6 (https://automeris.io/WebPlotDigitizer/): figure 1 from the study by Caravaggio et al [15] and supplementary figure 5 from the study by Dunn et al [16]. Original subject-wise data matrix was used for one study [17]. Subsequently, the standardized mean difference (Hedge’s g) of DR availability was calculated for each study.

**Figure 1.**
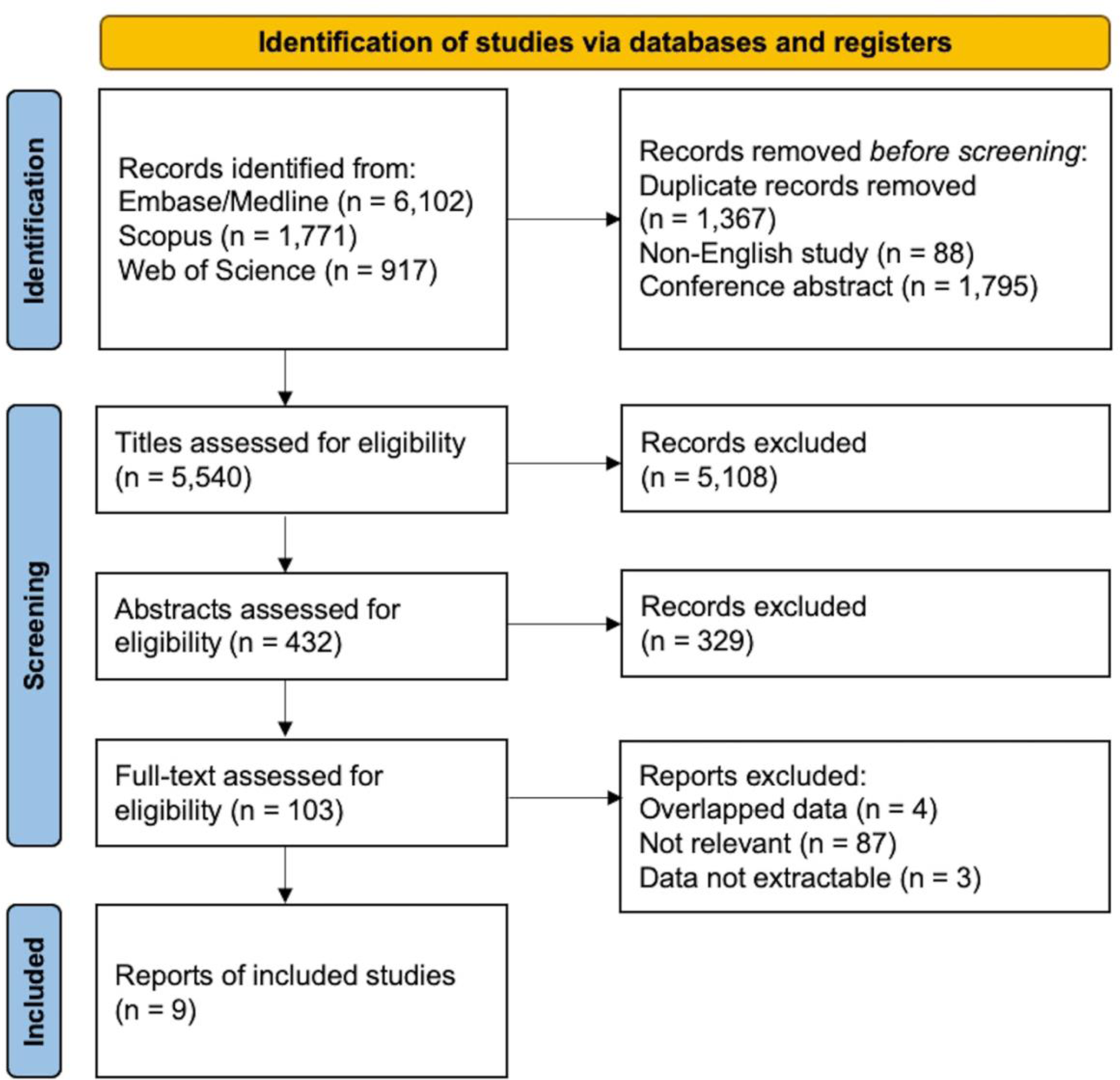
Flowchart for literature searches and data extraction

The standardized mean difference of DR availability between lean and overweight/obese subjects were first investigated using Bayesian hierarchical modelling with brms [18-20] that applies the Markov-Chain Monte Carlo sampling tools of RStan [21]. We set up a model with the standardized mean difference of DR availability as a dependent variable and added study, radiopharmaceutical and ROI as random intercepts to allow the standardized mean difference of DR availability to vary between studies, radiopharmaceuticals and ROIs. Bayesian models were estimated using four Markov chains, each of which had 4,000 iterations including 1,000 warm-ups, thus totaling 12,000 post-warmup samples. The sampling parameters were slightly modified to facilitate convergence (max treedepth=30). Also, complementary frequentist meta-analysis was performed using metafor [22]. Heterogeneity among studies was assessed using Cochran’s *Q* and *I^2^* statistics, as described previously [23]. The pooled effect size was estimated using random-effects model. Meta-regression was performed to explore potential sources of heterogeneity due to radiopharmaceutical and the mean BMI of overweight/obese subjects. Statistical analysis was carried out in R Statistical Software version 4.2.2 (The R Foundation for Statistical Computing).

## RESULTS

### Study Characteristics

The search identified 6,102 articles from Embase/Medline, 1,771 from Scopus, 917 from Web of Science. After excluding duplicate records (n=1,367), conference abstracts (n=1,795) and non-English publications (n=88), studies were assessed for eligibility after screening the title or abstract.

After reviewing the full text of 103 articles, nine studies were eligible for inclusion in this study [10-12, 15-17, 24-26] (Figure 1). Three studies with C11-raclopride [10, 17, 25], one with C11-PNHO [11], two with F18-fallypride [16, 24], one with I123-IBZM [26], one with C11-NMB [12] and one with both C11-raclopride and C11-PNHO [15] were included. The definition of lean and overweight/obese subjects varied across the studies. In 4 studies, subjects were divided into 2 groups according to BMI cutoff of 30 kg/m^2^ [10, 12, 17, 24]. Three studies that defined lean subjects as BMI less than 25 kg/m^2^ and obese subjects as BMI more than 30 [11, 16] or 35 kg/m^2^ [17] were included in both analyses of studies with BMI cutoff of 25 and 30 kg/m^2^. In one study, lean subjects were defined as BMI less than 24 kg/m^2^ and overweight subjects as BMI more than 27 kg/m^2^ [25]. As the study by Caravaggio et al included subjects of BMI between 18.6 and 27.8 kg/m^2^, we divided subjects into 2 groups with BMI cutoff of 25 kg/m^2^ [15]. Due to the heterogeneity in the definition of obesity, we ran the meta-analyses separately with datasets where obesity was defined as BMI > 25 kg/m^2^ or BMI > 30 kg/m^2^.

#### A. Brain Dopamine Receptor and Overweight/Obesity: A Bayesian Meta-Analysis

The distribution of the standardized mean difference of DR availability and mean BMI of overweight/obese subjects are shown in Figure 2 (BMI cutoff for overweight/obesity 25 kg/m^2^) and Figure 3 (BMI cutoff for obesity 30 kg/m^2^). In the analysis of BMI cutoff of 25 kg/m^2^, the standardized mean difference of DR availability between lean and obese subjects overlapped with zero. There was a more support for the association with radiopharmaceutical that overweight/obese subjects in studies with F18-Fallypride and C11-PHNO has higher DR availability and those in studies with C11-Raclopride and I123-IBZM has lower DR availability than lean subjects (Figure 2). In the analysis of BMI cutoff of 30 kg/m^2^, the standardized mean difference of DR availability between lean and overweight/obese subjects markedly overlapped with zero (Figure 3).

**Figure 2.**
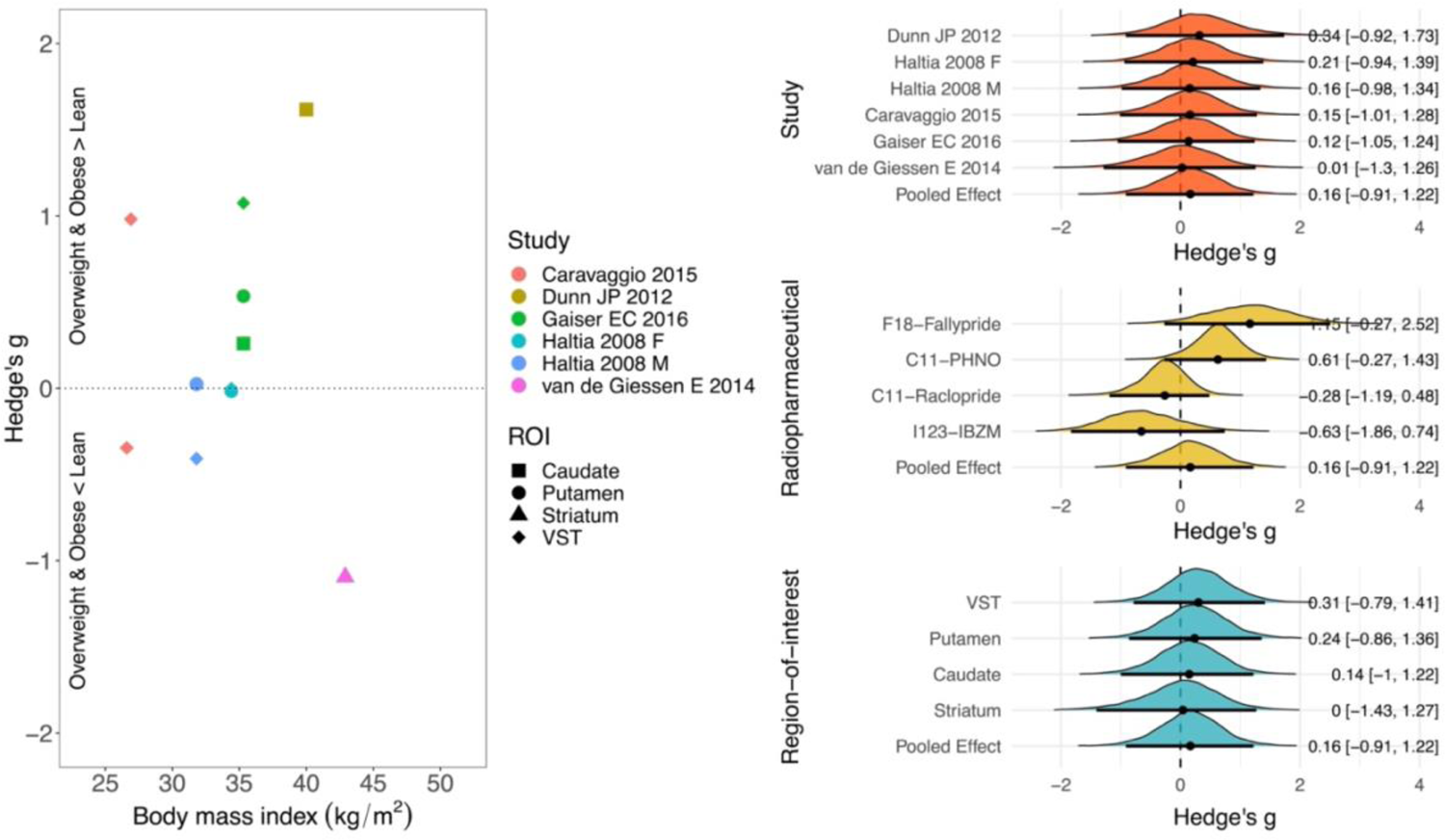
Studies of BMI cutoff of 25kg/m^2^, Left; scatter plot of the standardized mean difference of DR availability between lean and overweight/obese subjects and BMI, Right; Posterior probability distributions with their median (point), 80% (thick line) and 95% (thin line) posterior intervals, describing the standardized mean difference of DR availability. Posterior located on the positive side of the zero line suggests higher DR availability in overweight/obese subjects.

**Figure 3.**
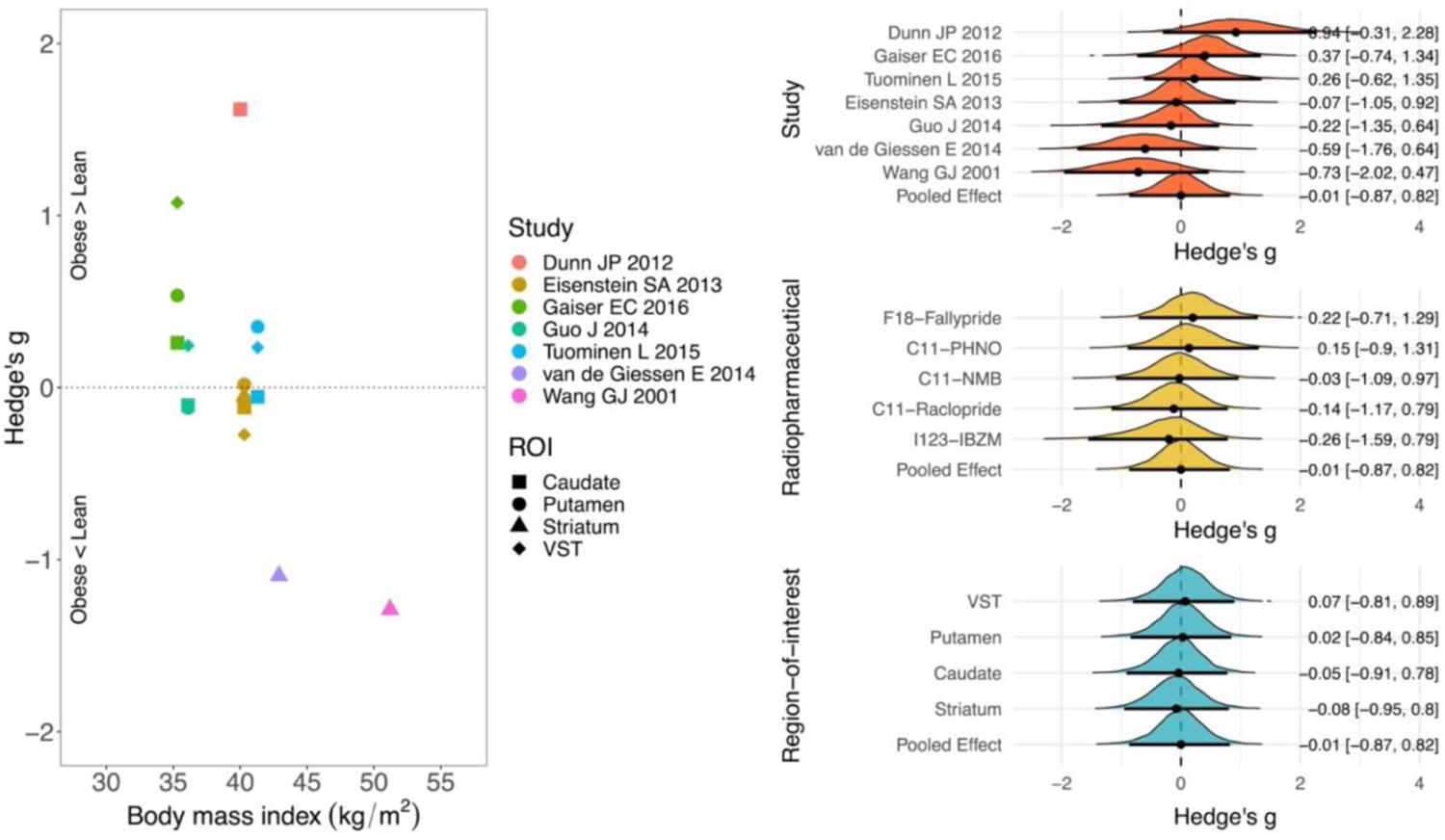
Studies of BMI cutoff of 30kg/m^2^, Left; scatter plot of the standardized mean difference of DR availability between lean and obese subjects and BMI, Right; Posterior probability distributions with their median (point), 80% (thick line) and 95% (thin line) posterior intervals, describing the standardized mean difference of DR availability. Posterior located on the positive side of the zero line suggests higher DR availability in obese subjects.

#### B. Brain Dopamine Receptor and Overweight/Obesity: Frequentist Meta-Analysis

A single ROI from each study was included in conventional meta-analysis. If the study reported results for more than one ROI [11, 17, 24, 25], mean DR availability across striatal subregions was calculated. In the meta-analysis of studies with BMI cutoff of 25 kg/m^2^, the pooled standardized mean difference of DR availability did not show the significant difference between lean and overweight/obese subjects (0.23, 95% confidence interval −0.46 ∼ 0.92, I^2^=76.7%). Meta-regression analyses revealed a statistically significant effect of radiopharmaceutical on the pooled effect size (C11-Raclopride, −1.0504, −1.8265 ∼ −0.2742, p=0.0080; I123-IBZM, −1.9141, −2.8656 ∼ −0.9626, p<0.0001). In a meta-analysis of studies with BMI cutoff of 30 kg/m^2^, the pooled standardized mean difference of DR availability was not significantly different between lean and overweight/obese subjects (0, −0.69 ∼ 0.69, I^2^=83.4%). Meta-regression analyses revealed that the mean BMI of overweight/obese subjects is negatively associated with the standardized mean difference of DR availability (−0.1214, −0.2397 ∼ −0.0031, p=0.0442) (Figure 4).

**Figure 4.**
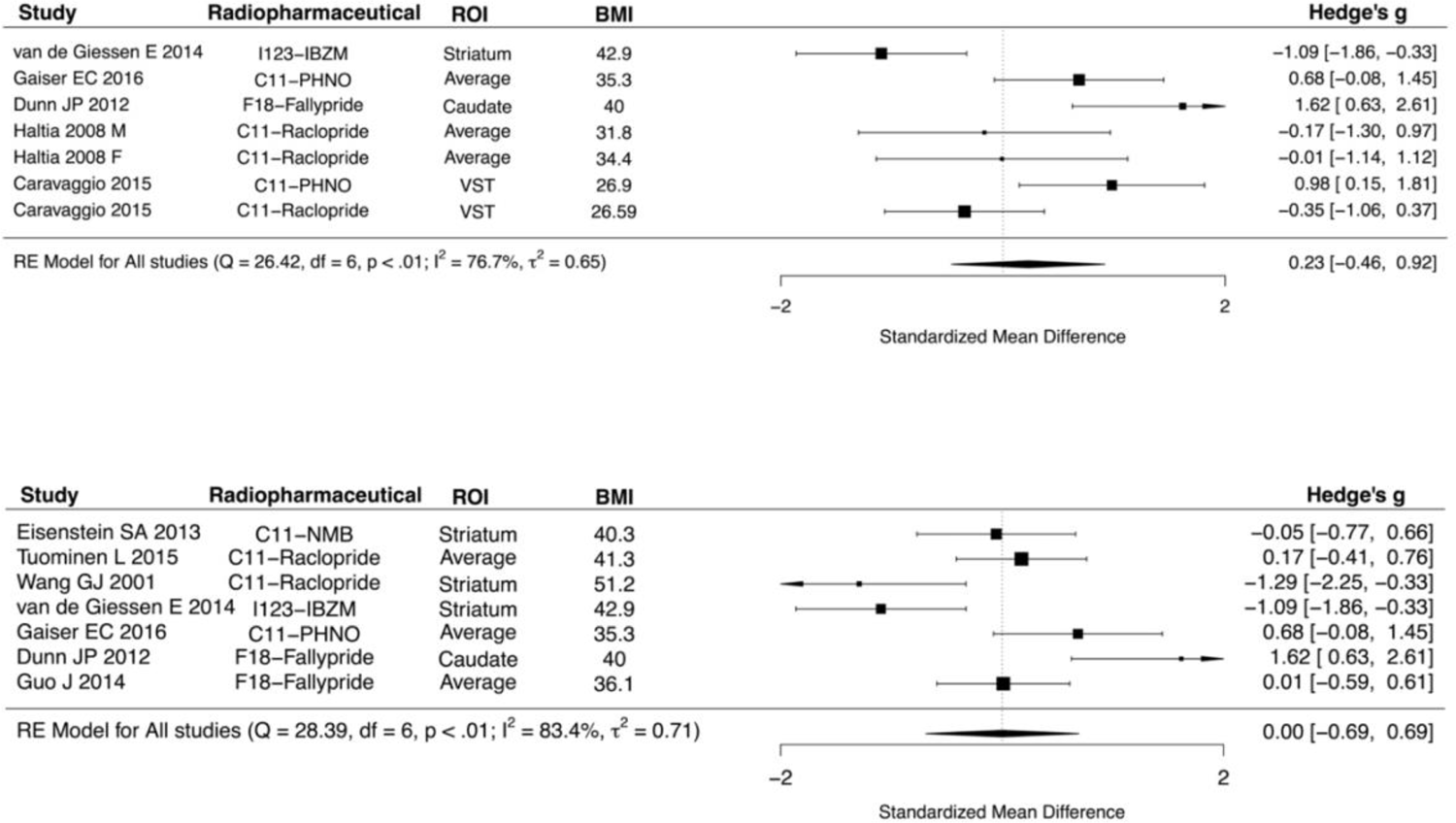
Forest plots of studies of BMI cutoff of 25 and 30kg/m^2^

## DISCUSSION

Our main finding was that brain DR availability does not differ between lean and overweight/obese subjects. This result was confirmed in both Bayesian and conventional meta-analyses and with two different criteria (BMI > 25 kg/m^2^ or BMI > 30 kg/m^2^) for overweight/obesity. However, the effect sized varied as the function of the used radiopharmaceuticals and the degree of obesity. Altogether these results suggest that striatal downregulation of D2R is not a common feature of obesity despite the centrality of dopaminergic system in feeding and reward [27].

Obesity is caused by an imbalance between the energy intake and expenditure [28]. Feeding is controlled by both homeostatic regulatory brain circuits and those involved in reward and motivation [29]. Dopamine is one of the neurotransmitters involved with eating behavior through modulation of the rewarding properties of food and the motivation and desire for food consumption [27]. There are two major hypotheses regarding the role of dopamine in obesity. The first hypothesis, dopamine hyperresponsiveness, proposes that there is a hypersensitivity to rewards and increased behavioral salience toward food, resulting in the excessive intake of palatable foods [30, 31]. The second hypothesis, reward deficit model, states that subjects who are insensitive to rewards overeat to increase their endogenous dopamine levels to a normal amount of pleasure [30, 31]. However, it is not possible to measure the synaptic dopamine levels directly in the human brain. Therefore, PET with DR radiopharmaceuticals has been used to understand the role of dopaminergic system in obesity. However, it is difficult to interpret the relationships between DR availability measured from PET scans and the synaptic dopamine level. Low DR binding potential could represent 1) low density of existing DR, 2) low affinity to bind DR or 3) greater amount of endogenous dopamine which competes with DR radiopharmaceutical to bind DR [14, 24].

Nine such studies were included in this meta-analysis. There was no significant difference of DR availability between lean and overweight/obese subjects in 4 studies in any striatal ROIs [12, 17, 24, 25], in 1 study no effect in caudate and putamen [11] and also in 1 study with C11-Raclopride, there was no significant correlation between BMI and DR availability [15]. In the study by Dunn et al, obese subjects had higher DR availability in caudate [16] and in the study by Caravaggio et al with C11-PHNO, there was a positive correlation between BMI and DR availability [15]. In 2 studies, obese subjects had lower DR availability than lean subjects [26, 29].

Because the studies had different definition of overweight/obese subjects, we conducted two separate meta-analyses with studies using two different cutoffs for obesity (BMI > 25 kg/m^2^ and BMI > 30 kg/m^2^). Neither meta-analysis provided support for overall obesity-dependent modulation of striatal DR. In the meta-analysis of BMI cutoff of 25 kg/m^2^, 5 studies were included. The study by Caravaggio et al. included overweight subjects of BMI between 25 and 27.8 kg/m^2^ without obese subjects [15] and the study by Haltia et al. included both overweight and obese subjects from BMI 27 kg/m^2^ [25]. The other 3 studies were included in both analysis of BMI cutoff of 25 and 30 kg/m^2^ [11, 16, 26]. Although there was no difference between lean and overweight/obese subjects in DR availability, there was an effect of radiopharmaceutical on both Bayesian and conventional meta-analysis of BMI cutoff of 25 kg/m^2^. One study with F18-Fallypride [16] and 2 studies with C11-PHNO showed the higher DR availability in overweight/obese subjects while 2 studies with C11-Raclopride [15, 25] and 1 study with I123-IBZM [26] did not show the difference of DR availability, which might elucidate this effect of radiopharmaceutical on the result.

In a meta-analysis with BMI cutoff of 30 kg/m^2^, in addition to 3 studies that included in both analysis of BMI cutoff of 25 and 30 kg/m^2^, 4 additional studies with obese subjects were included. Similar with the result of BMI cutoff of 25 kg/m^2^, there was no difference between lean and overweight/obese subjects in DR availability. However, meta-regression showed that the mean BMI of overweight/obese subjects was negatively associated with the standardized mean difference of DR availability. The subjects in the studies with the two different BMI cutoffs also varied with respect to the degree of the severity of obesity: The mean BMI of overweight/obese subjects in a meta-analysis of BMI cutoff of 30 kg/m^2^ was higher than that of 25 kg/m^2^ (mean BMI of 40.1 kg/m^2^ vs 34.9 kg/m^2^). Also, the study by Wang et al [10] with the highest mean BMI of obese subjects (mean BMI of 51.2 kg/m^2^) was included in a meta-analysis of BMI cutoff of 30 kg/m^2^. The meta-analysis with BMI cutoff of 30 kg/m^2^ thus also includes obese subjects with the wider range of BMI.

Against our expectation, there was no effect of ROI on the standardized mean difference of DR availability between lean and overweight/obese subjects, although all ROIs could be included in Bayesian meta-analysis (unlike in the frequentist meta-analysis that included a single representative ROI from each study). Five DR radiopharmaceuticals were included in this meta-analysis and each radiopharmaceutical has its own profile of DR affinity, competition with endogenous dopamine and agonist/antagonist. Both C11-Raclopride and F18-Fallypride, D2/D3 antagonists, have a similar affinity to D2 and D3 and compete with endogenous dopamine [16, 32]. I123-IBZM, a D2/D3 antagonist for SPECT, competes with endogenous dopamine, similar with C11-Raclopride [14]. C11-NMB, a D2-selective antagonist, is not replaceable by endogenous dopamine [12]. C11-PHNO, a D3-preferring agonist, is more sensitive to endogenous dopamine levels than radiopharmaceuticals of DR antagonists [11], therefore, considered superior to detect synaptic dopamine release [33]. Therefore, it is critical to consider the feature of each radiopharmaceutical while interpreting the results from PET and SPECT scans.

There are several limitations in this study. First, only a small number of studies could be included in this meta-analysis. After a systematic search for publication, several papers were assessed for eligibility. However, most of them were published from the same institution with overlapping subjects and we selected the most relevant publication among the pool of articles with same subjects. In addition, as two different BMI cutoffs were applied in these studies, 5 studies with BMI cutoff of 25kg/m^2^ and 7 studies with that of 30 kg/m^2^ could be included in meta-analysis. Finally, five DR radiopharmaceuticals were used in studies included in this meta-analysis and each radiopharmaceutical has a its own receptor-binding property. Therefore, we should be cautious when interpreting the findings from the studies included in this meta-analysis.

In conclusion, brain DR is not different between lean and overweight/obese subjects and these meta-analytic findings from patients also align with the recent large-scale study of non-obese healthy subjects [34]. There is an effect of the variety of radiopharmaceuticals and the degree of the severity of obesity on this result, therefore, we still cannot exclude the association of brain DR with obesity.

For example, it is possible that obesity alters the threshold for endogenous dopamine release following feeding rather than the receptor densities [8], or that obesity alters the molecular coupling between dopamine and other neurotransmitter systems such as endogenous opioid systems that has been consistently linked with obesity [35-37]. Further studies with multi-radiopharmaceutical in the same individuals are needed to elucidate the potential links between DR and obesity.

## DATA AVAILABILITY

The datasets generated during and/or analysed during the current study are available from the corresponding author on reasonable request

## Data Availability

All data produced in the present study are available upon reasonable request to the authors

## TABLES

**Table 1.**
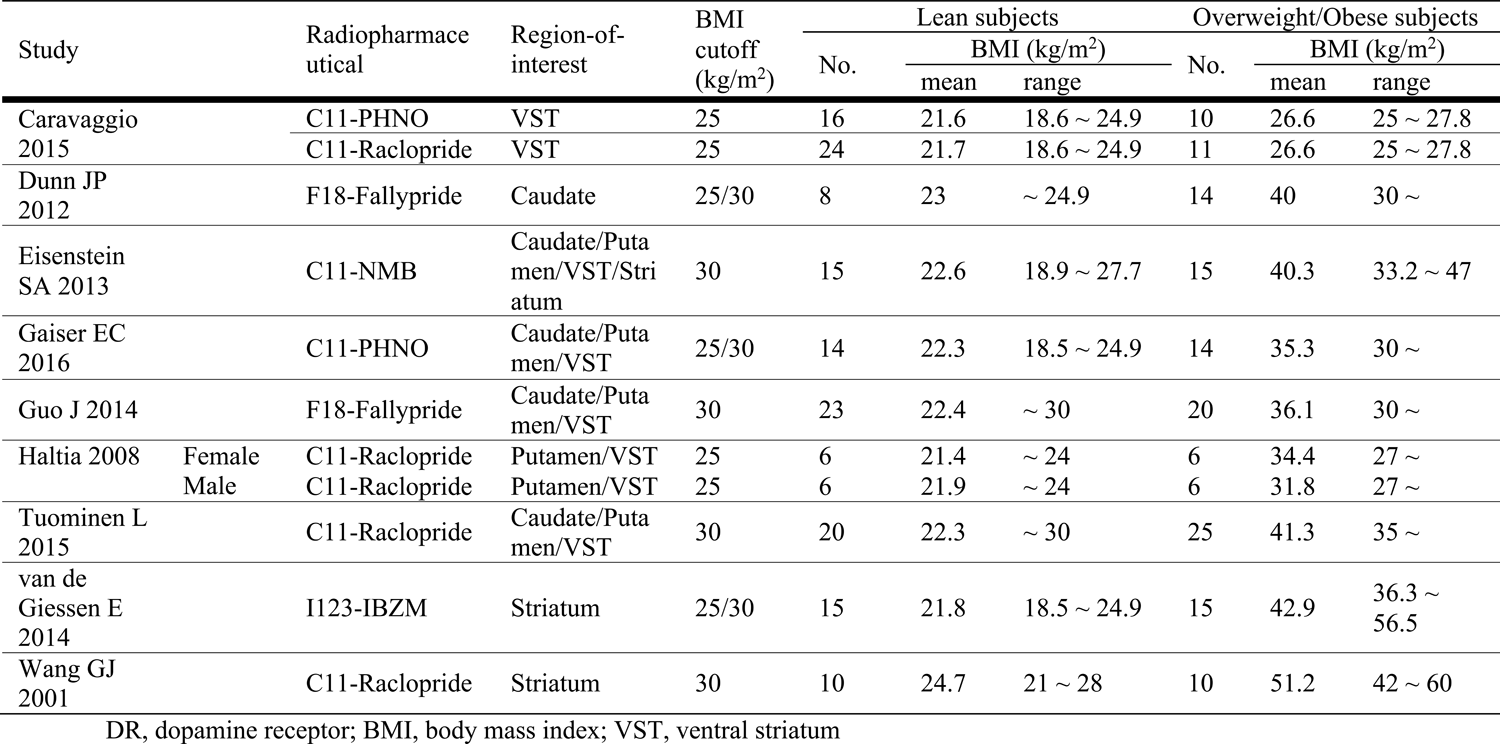
Studies of DR availability

